# Effectiveness and Tolerability of Nonmedical Switching from Originator (MabThera®) to Biosimilar (Truxima®) Rituximab in People with Multiple Sclerosis: A Tertiary Single-Center Observational Study

**DOI:** 10.64898/2026.08.01.26359456

**Authors:** Ahmed H. Althobaiti, Afnan A. Alnughaimish, Saif S. Alqahtani, Fahad Aldosari

## Abstract

**Background:** Rituximab is used off-label for multiple sclerosis (MS), and biosimilar substitution raises a distinct extrapolation challenge, as MS is not an approved indication for the reference product. Real-world nonmedical-switching data inform biosimilar appropriateness decisions by clinicians, societies, and payers.

**Objective:** To report the effectiveness and tolerability of nonmedical switching from originator (MabThera®) to biosimilar rituximab (Truxima®) in people with MS (pwMS).

**Methods:** A retrospective, single-center observational cohort study of 50 pwMS switched after at least two originator infusions, followed for two years.

**Results:** Annualized relapse rate declined from 0.45 (95% CI 0.28–0.68) pre-rituximab to 0.02 (95% CI 0.00–0.13) on originator and 0.00 (95% CI 0.00–0.05) on biosimilar (p = 0.367 between products). In paired imaging analysis (n = 29), the proportion with active scans declined progressively (50.0%, 34.5%, 17.2%; Cochran’s Q, p = 0.040), with no difference between the originator and biosimilar periods (McNemar, p = 0.227). B-cell depletion deepened progressively. All patients remained on biosimilar through the end of follow-up.

**Conclusion:** Nonmedical switching from originator to biosimilar rituximab was associated with comparable clinical and radiological outcomes, supporting its use in pwMS without concern for inferior efficacy or diminished tolerability.

## Introduction

Rituximab demonstrated efficacy in MS in the phase 2 HERMES trial (14.5% vs. 34.3% relapse-free vs. placebo at week 24; 91% reduction in contrast-enhancing lesions vs. placebo)^1^, and later showed superiority over dimethyl fumarate in the RIFUND-MS phase 3 trial^2^. Despite this growing body of trial and real-world evidence, rituximab remains off-label in MS, lacking regulatory approval for the indication^2,3^.

Truxima® (rituximab-abbs, Celltrion Inc.) is, a biosimilar of the reference monoclonal antibody MabThera® (rituximab, Genentech Inc.), approved by the the EMA in April 2017 and by the FDA in November 2018.^4,5^.

Physician skepticism toward biosimilars often centers on extrapolation — the regulatory mechanism by which approval in one indication is extended to others based on shared biology and mechanism of action.^1^ Extrapolation is not unique to biosimilars — it also underpins the continued use of originator biologics, which undergo manufacturing changes over time that alter the product itself^7^ — and is justified when comprehensive comparability analysis shows the mechanism of action is consistent across indications^8^.

Biosimilar approval rests on analytical characterization, in vitro functional assays, and PK/PD studies that differ fundamentally from the clinical trial evidence physicians are trained to assess. In a survey by Cohen et al., only half of respondents were familiar with this “totality of evidence” framework, and just 12% were comfortable with extrapolation^9^. Several learned societies have discouraged biosimilar use in extrapolated indications^10–14^, and half of physicians do not recommend switching to a biosimilar in stable patients^15^ — though a separate review found no evidence of patient harm^16^.

This is particularly relevant for rituximab, where MS is not an approved indication even for the reference product — placing it beyond the scope of standard extrapolation frameworks, which extend approval only across indications already held by the originator. In this setting, biosimilar appropriateness is not determined by direct regulatory judgment but by clinicians, scientific societies, and payers drawing on multiple lines of evidence, including real-world outcomes^17^. Real-world data on nonmedical switching from originator to biosimilar rituximab in pwMS are lacking.

Here we report the effectiveness and tolerability of nonmedical switching from originator rituximab (MabThera®) to biosimilar rituximab (Truxima®) in pwMS over approximately two years of follow-up.

## Methods

### Study design and population

This was a retrospective observational cohort study of pwMS diagnosed by 2017 McDonald criteria who received at least two originator rituximab (MabThera®) infusions followed by a nonmedical switch to biosimilar rituximab (Truxima®) at King Saud Medical City, Riyadh, Saudi Arabia. Patients were enrolled by consecutive sampling; originator treatment began between December 2015 and April 2022. Induction was two 1,000 mg infusions on days 0 and 14, with maintenance every six months, dosed by body surface area. Follow-up was truncated at two years from the first biosimilar infusion, since data completeness was validated only to that point. The study was approved by the Institutional Review Board (H1RI-04-Apr23-04); informed consent was waived given exclusive reliance on electronic medical records.

### Statistical analysis

Analyses were performed in Python 3.12.12 (pandas 3.0.1, NumPy 2.4.2, SciPy 1.17.1, matplotlib 3.10.8, statsmodels 0.14.6, pingouin 0.6.1). Continuous variables were assessed for normality (Shapiro–Wilk, n 50; Kolmogorov–Smirnov, n > 50) and reported as mean ± SD or median (IQR); categorical variables are presented as counts with percentages. Variables available in 60% of the relevant unit of analysis (patients, scans, visits, or labs) were included in the primary analysis; others are reported in the supplementary material.

### Outcomes

#### Annualized relapse rate

Patients were scheduled for follow-up every six months, assessed throughout by the same MS specialist (A.H.A.). At each visit, patients were assessed for relapses - new neurological symptoms lasting 24 hours without fever or infection with new objective findings on examination -; concomitant imaging activity was not required^18^. ARR was calculated across three periods: (i) pre-rituximab, the year before starting rituximab; (ii) originator, from first rituximab infusion to the day before first biosimilar administration; and (iii) biosimilar, from first biosimilar infusion to last follow-up. All visits, scans, and labs were assigned to periods using this date-based rule, and ARR was derived per period as total relapses divided by total patient-years. Poisson regression with log-transformed exposure as an offset term was used for between-period comparison, substituting negative binomial regression if the deviance-to-degrees-of-freedom ratio exceeded 1.5; zero-event periods used one-sided 95% exact Poisson confidence intervals and mid-p exact pairwise tests.

#### Disability progression

For each disability measure Expanded Disability Status Scale, Nine-Hole Peg test, and Timed 25-Foot Walk (EDSS, NHPT, T25FW), baseline was the value at the initial visit or, if unavailable, the first available follow-up score. Patients were eligible for progression analysis with a baseline score and at least two follow-up assessments 6 months apart. EDSS worsening was defined as an increase of 1.5 points from a baseline of 0, 1.0 from a baseline >0 and <5.5, or 0.5 from a baseline 5.5; NHPT and T25FW worsening was a 20% increase from baseline. Each event was classified as relapse-associated worsening or progression independent of relapse biology (PIRB). Between-product comparisons used the Fisher exact test.

#### Treatment persistence

The proportion of patients remaining on biosimilar treatment through the end of follow-up was reported descriptively.

#### Radiological activity

Radiological activity was defined as any new or enlarging T2 lesion, or contrast-enhancing lesion (CEL), on brain or spinal cord imaging across the originator and biosimilar periods; the pre-rituximab baseline used CELs only, as most patients lacked a prior scan. Patients could have zero or more scans per period; at the patient level, scans within a period were collapsed to a single binary (active/inactive) outcome, and at the scan level the proportion of active scans per period was reported. The primary analysis compared the proportion of active patients across periods by chi-square test with pairwise Fisher exact tests (Bonferroni = 0.0167). A paired analysis (n = 29 with data across all three periods) used Cochran’s Q test with post-hoc McNemar tests for the binary outcome, and the Friedman test with post-hoc Wilcoxon signed-rank tests for active-scan proportions, both Bonferroni-corrected. Stratification by exposure duration (12 vs. >12 months) is reported in the Supplementary Material.

#### B-cell depletion

Two B-cell monitoring tests were available as part of routine care, at the treating physician’s discretion: a lymphocyte flow cytometry panel reporting absolute counts and percentages of T, B, and NK cells, and a separate semi-quantitative CD19/CD20 percentage assay; full assay protocols are in the supplementary material. B-cell depletion was defined as CD19/CD20 percentage <1% or absolute CD19 count <10 cells/µl. Laboratory tests were assigned to periods using the same date-based rule, with each patient’s observations within a period collapsed to a single median value. The paired analysis used the Friedman test with post-hoc Wilcoxon signed-rank tests; the unpaired analysis used the Kruskal–Wallis test with post-hoc Mann–Whitney U tests; both were Bonferroni-corrected, with effect sizes expressed as the rank-biserial correlation coefficient (r). The interval between each test and the preceding infusion was compared between periods using the Mann–Whitney U test.

## Results

### Characteristics of the population

Fifty pwMS diagnosed by 2017 McDonald criteria were included, all receiving both reference and biosimilar products; baseline characteristics are in Table 1.

**Table 1.** Baseline Characteristics of the Study Cohort (N = 50)

| Characteristic | All patients (N = 50) |
| --- | --- |
| Age, years | 27 (24–33) |
| Sex, female | 38 (76.0%) |
| Disease duration, years | 1 (0–4) |
| EDSS at baseline | 1 (1–3.5) |
| Relapses 1 year prior to rituximab | 0 (0–1) |
| Patients with 1 relapse, 1 year prior | 21 (42.9%) |
| ARR, 1 year prior | 0.45 (95% CI 0.28–0.68) |
| T2 lesion burden on pre-rituximab MRI |  |
| — >20 lesions | 37 (74.0%) |
| — 10–20 lesions | 8 (16.0%) |
| — 1–9 lesions | 5 (10.0%) |
| Gadolinium-enhancing lesions | 19 (40.4%) |
| Median CELs among those with enhancement | 1 (0–2) |
| Previous DMT |  |
| — Treatment-naïve | 21 (42.0%) |
| — Natalizumab | 12 (24.0%) |
| — Fingolimod | 6 (12.0%) |
| — Interferon beta-1b (Betaferon) | 5 (10.0%) |
| — Interferon beta-1a IM (Avonex) | 3 (6.0%) |
| — Teriflunomide | 2 (4.0%) |
| — Interferon beta-1a SC (Rebif) | 1 (2.0%) |
| Baseline CD19 absolute count, cells/μl | 272.6 (171.5–394.2) |
| Baseline CD19/CD20 percentage | 13.10% (SD 6.27) |

Data are median (IQR) or n (%) unless otherwise stated.
ARR = annualized relapse rate; CEL = contrast-enhancing lesion; DMT = disease-modifying therapy; EDSS = Expanded Disability Status Scale; IQR = interquartile range; SD = standard deviation.
Available in 43/50 (86.0%) patients
Available in 49/50 (98.0%) patients; 1 patient had no documented visit in the year prior
Available in 47/50 (94.0%) patients; 3 patients did not receive gadolinium
Available in 33/50 (66.0%) patients
Available in 35/50 (70.0%) patients

### Outcome measure differences between products

Substantial asymmetries in data collection were observed between the originator and biosimilar treatment periods, including shorter follow-up duration, fewer scans, and fewer laboratory tests during the originator period (Supplementary Table S2).

### Clinical outcomes

Throughout the study period, 256 clinical follow-up visits occurred (relapse status not recorded at 6 visits), with a median of 5 visits per patient (IQR 4–6) over a median total follow-up of 31.3 months (IQR 26.3–33.8). One relapse occurred during the originator period, 8.3 months after induction; no relapses occurred during the biosimilar period, over 75.6 patient-years. The ARR was 0.45 (95% CI 0.28–0.68) prior to rituximab, 0.02 (95% CI 0.00–0.13) during the originator period, and 0.00 (95% CI 0.00–0.05, upper bound only) during the biosimilar period.

Compared with the pre-rituximab period, the ARR was significantly reduced during both the originator (94.9% reduction; exact conditional Poisson, p < 0.001) and biosimilar periods (100% reduction; Fisher exact, p < 0.001), with no significant difference between originator and biosimilar periods (Fisher exact, p = 0.367).

**Figure 1.**
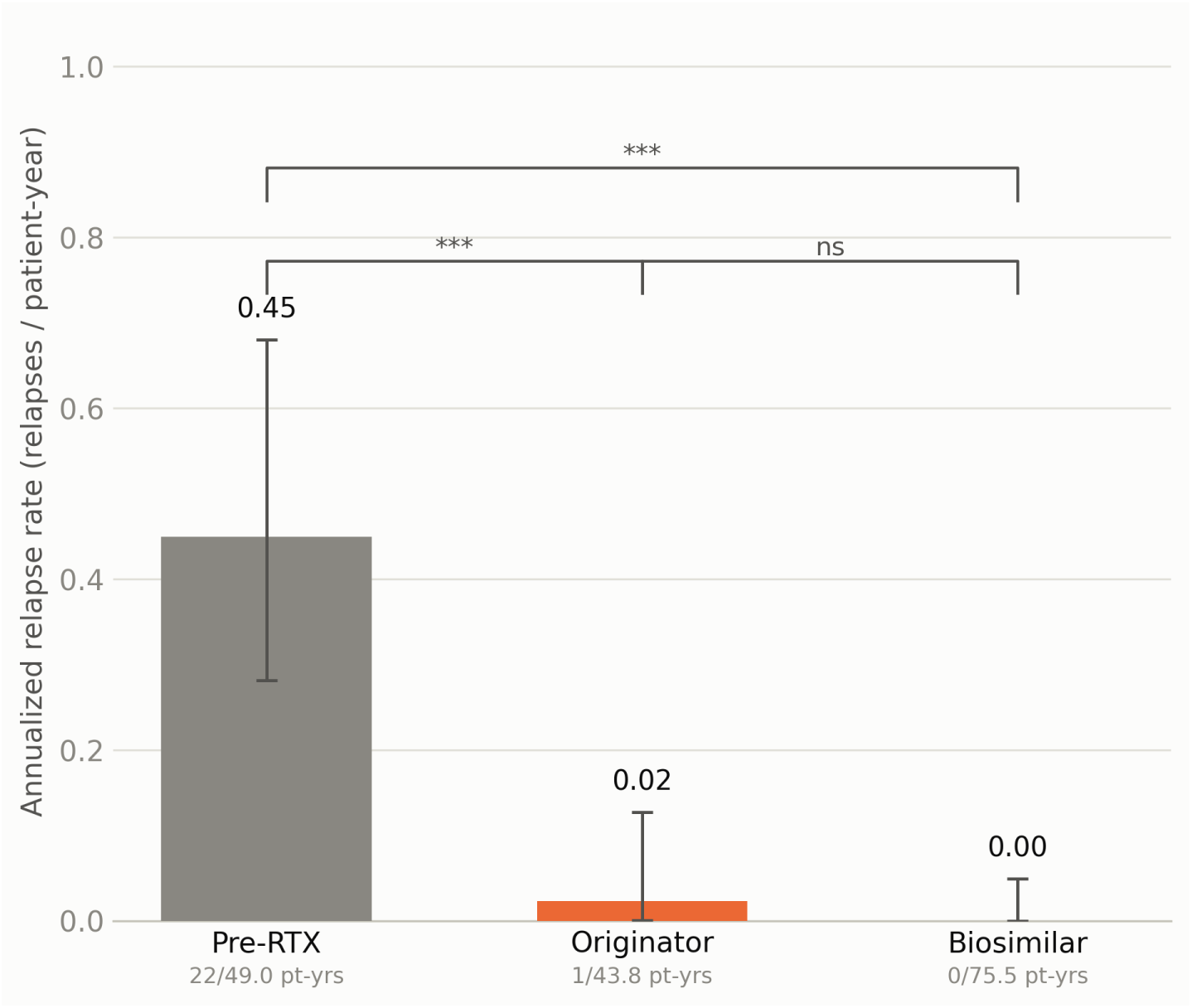
Annualized relapse rate across treatment eras. ARR = events / patient-years; 95% CI shown as error bars (exact Poisson; one-sided upper bound where events = 0). p-values: exact conditional Poisson test, or mid-p exact / Fisher’s exact when either group has 0 events. ns = not significant (p 0.05).

Because few patients met the eligibility criteria defined in the methods, disability progression analysis is reported in the supplementary material (Supplementary Table S1).

All patients remained on biosimilar treatment through the end of follow-up, approximately two years after the first biosimilar switch.

### Radiological outcomes

A total of 103 MRI scans were performed (45 originator-period, 58 biosimilar-period), with at least one scan available for 36 patients during the originator period and 40 during the biosimilar period; 29 patients had scans across all three periods, eligible for paired analysis. Scan timing and interval data are in the supplementary material (Supplementary Table S2).

Among the 29 patients with MRI data across all three periods, the proportion with active disease declined from 14 of 28 (50.0%; one patient had a non-contrast pre-rituximab study and was excluded from that period’s denominator) pre-rituximab to 10 of 29 (34.5%) during the originator period and 5 of 29 (17.2%) during the biosimilar period — significant across periods by both binary patient-level classification (Cochran’s Q = 6.42, p = 0.040) and active-scan proportion (Friedman ² = 8.67, p = 0.013). Pairwise comparisons with Bonferroni correction (= 0.0167) identified the pre-rituximab period as the principal driver: pre-rituximab vs. biosimilar reached significance on the Wilcoxon test (median 0.50 vs. 0.00; W = 13.0, p = 0.002) but not McNemar (p = 0.035), while originator vs. biosimilar reached significance on neither McNemar (p = 0.227) nor Wilcoxon (W = 16.0, p = 0.064).

Analysis stratified by time for the full cohort is provided in the Supplementary Material (Supplementary Tables S3–S4, Supplementary Fig 1).

### CD19/CD20 laboratory outcomes

Blood samples for CD19/CD20 testing were obtained from 35 patients prior to rituximab initiation, 42 patients during the originator period, and 44 patients during the biosimilar period, yielding 35, 78, and 100 samples respectively.

Absolute CD19 count declined progressively across periods, from a median of 272.6 cells/µl prior to rituximab to 64.6 cells/µl during the originator period and 14.9 cells/µl during the biosimilar period. Using a depletion threshold of <10 cells/µl, depletion rates were 5 of 33 (15.2%) prior to rituximab, 13 of 25 (52.0%) during the originator period, and 23 of 26 (88.5%) during the biosimilar period — a significant difference across periods (Kruskal–Wallis H = 32.32, p < 0.001), with pairwise Mann–Whitney U tests (Bonferroni = 0.0167) confirming a significant originator-vs-biosimilar difference (U = 453.0, p = 0.016). The unpaired analysis is reported here; a paired analysis (n = 7) is provided in the Supplementary Material (Supplementary Figure 5).

CD19/CD20 percentage followed a similar pattern, declining from a mean of 13.10 ± 6.27% prior to rituximab (normally distributed) to a median of 1.00% (IQR 0.10–5.20) during the originator period and 0.66% (IQR 0.00–2.98) during the biosimilar period (non-normally distributed thereafter). Using a depletion threshold of 1%, depletion rates were 0 of 35 (0.0%) prior to rituximab, 34 of 65 (52.3%) during the originator period, and 55 of 100 (55.0%) during the biosimilar period — significant across periods (Kruskal–Wallis H = 66.59, p < 0.001), though the originator-vs-biosimilar difference did not reach significance after Bonferroni correction (Mann–Whitney U = 3757.5, p = 0.087). The unpaired analysis is reported here; a paired analysis (n = 26) is provided in the Supplementary Material (Supplementary Figure 5).

**Figure 2.**
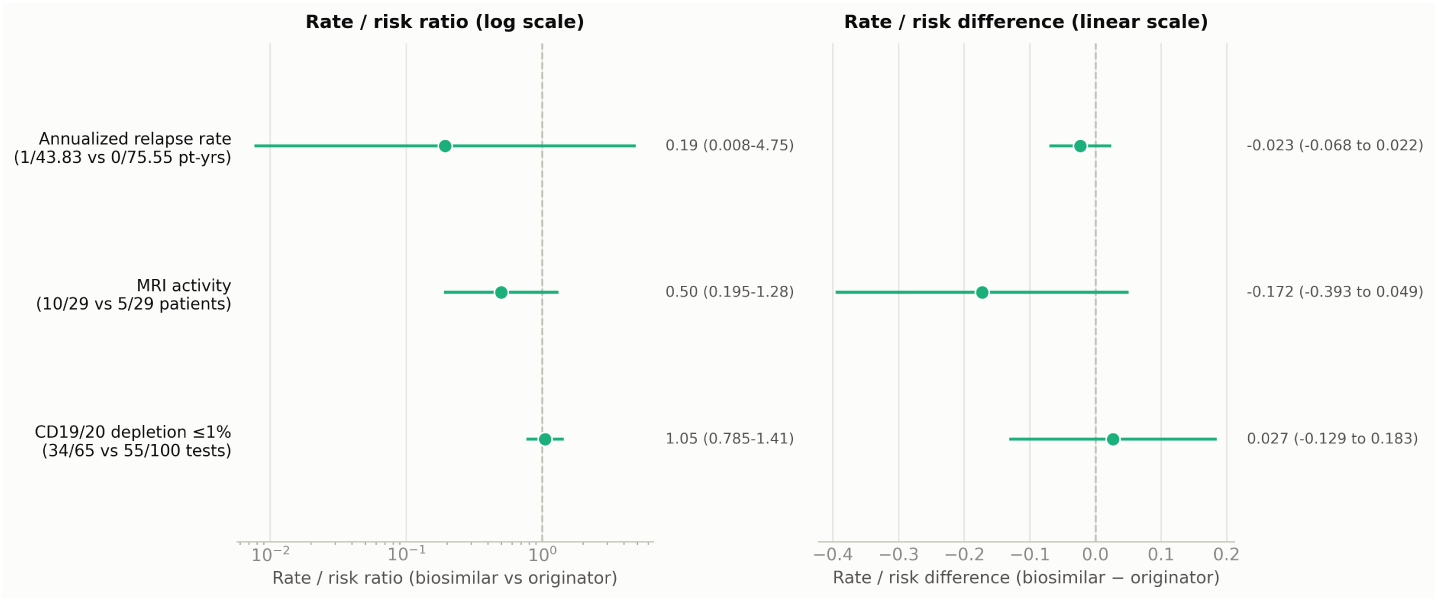
Originator vs biosimilar rituximab: comparative effect estimates. Rate/risk ratio and difference use Wald (log-scale / linear) 95% CIs on independent groups. ARR ratio uses a 0.5 continuity correction (zero relapse events observed on biosimilar) — wide, descriptive CI only. Dashed line = no difference (ratio = 1, difference = 0).

**Figure 3.**
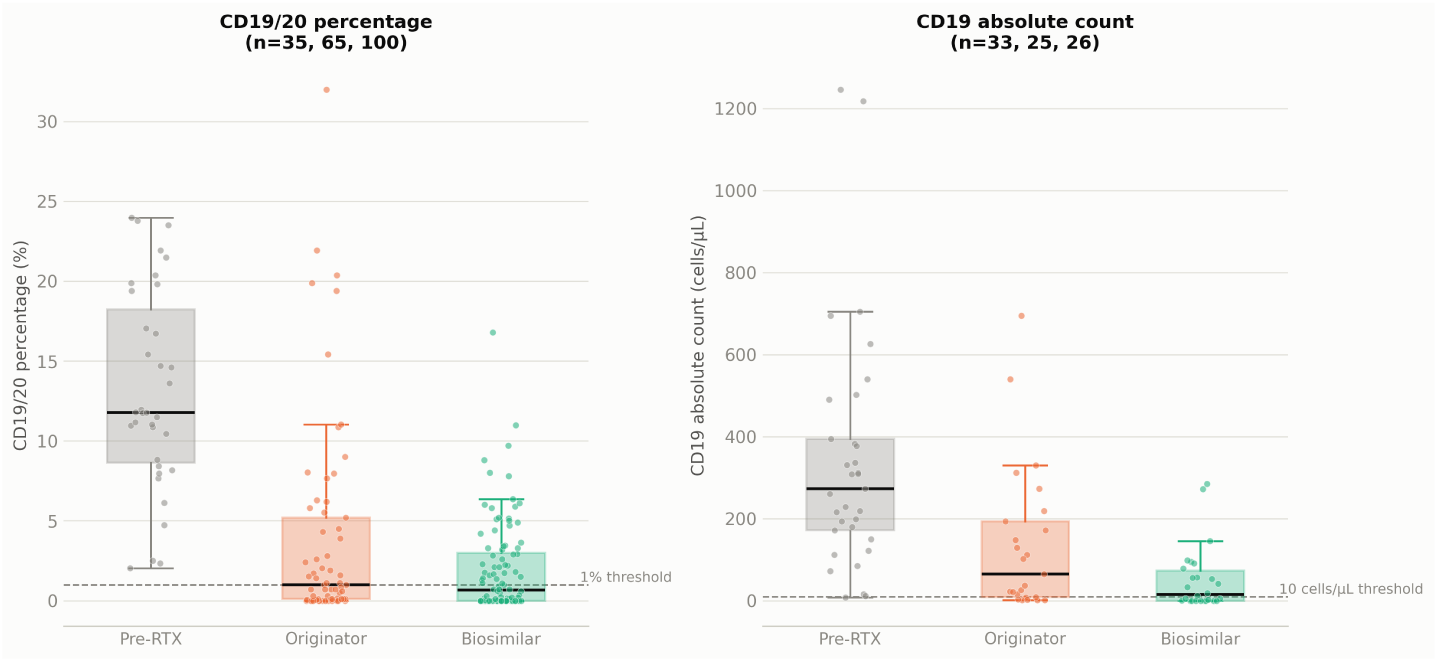
CD19/20 B-cell measures across treatment eras (all available tests, unpaired). Boxes: median and IQR; whiskers extend to 1.5×IQR. Points are individual test results, jittered horizontally for visibility. Not all patients contribute to every era.

## Discussion

To our knowledge, this is the first study to report on the effectiveness of nonmedical switching from originator rituximab (MabThera®) to biosimilar rituximab (Truxima®) in pwMS. In this real-world cohort, both products were associated with substantial and comparable control of relapses.

Radiological outcomes followed a similar trajectory but require closer examination of timing. Most originator-period scans were obtained within the first 12 months of treatment, a window during which B-cell-depleting therapy has not yet achieved its full suppressive effect on radiological activity — likely explaining why the originator period, taken as a whole, did not differ significantly from the pre-rituximab baseline. When stratified by duration of exposure, however, a clearer pattern emerged: activity on scans obtained beyond 12 months of originator treatment was comparable to activity within the first 12 months of biosimilar treatment, and continued to decline further beyond 12 months of biosimilar treatment (Supplementary Material: Supplementary Figure 1).

All patients remained on biosimilar treatment without discontinuation through the end of follow-up, approximately two years after the first biosimilar switch. Although tolerability and infusion-related adverse events were not systematically assessed, this complete persistence — together with the absence of any documented safety signal on retrospective chart review — suggests the switch was well tolerated in clinical practice.

A deeper peripheral B-cell depletion was observed with the biosimilar than with the originator — yet without additional clinical or radiological benefit, raising the question of whether rituximab’s MS effect is fully explained by CD20^+^ B-cell depletion. Extrapolation assumes the mechanism driving efficacy in the approved indication also drives efficacy in the extrapolated one. For rituximab in MS, this is vulnerable: biosimilar comparability rests primarily on antigen-antibody binding and CD20^+^ depletion, yet alternative mechanisms have been proposed — effects on CD3^+^CD20^+^ cells, PD-L1-expressing monocytes, antigen-specific CD8^+^ T cells, and lymphocyte adhesion/activation networks — any of which could let a biosimilar pass every regulatory metric while differing from the originator in a dimension never evaluated.

This concern is reinforced by the MS-specific discordance between depletion and efficacy: patients often remain stable despite variable depletion/repletion, while breakthrough activity despite adequate depletion is uncommon and concentrated early in treatment — consistent with our cohort^19,20^. This contrasts with other rituximab-treated diseases, where repletion does track relapse: neuromyelitis optica spectrum disorder (NMOSD)^21,22^, rheumatoid arthritis^23,24^, and pemphigus vulgaris^25,26^.

This uncertainty is reflected in real regulatory divergence: Truxima received EMA approval for all originator indications, including pemphigus vulgaris^4^, but FDA approval excluded pemphigus vulgaris despite the originator itself holding that approval in the US^5^. This shows extrapolation judgments are not settled by consensus, even for the same biosimilar against the same originator. Real-world data carry strong evidentiary weight when extrapolation involves a complex or incompletely understood mechanism, or when patient/disease characteristics differ substantially from the approved indication — both true here, given the originator itself was never approved for MS. The question is therefore not only whether the biosimilar behaves like the originator, but whether the originator’s off-label evidence in MS extends to the biosimilar. No biosimilar has been granted approval for an indication the reference product itself never held, and proposed frameworks for reasoning through such questions have come from individual stakeholders, not regulators^17^.

This off-label extrapolation problem doesn’t arise for biosimilars of originators already approved for MS. Their approval rests on conventional analytical comparability and the standard regulatory framework of totality of evidence, further reinforced by phase 3 trial data for both Tyruko® (biosimilar natalizumab)^27^ and Xacrel (biosimilar ocrelizumab)^28^.

Our findings can be considered alongside Perez et al.^29^, whose between-group design compared originator and biosimilar in two separate groups (105 vs. 40) allocated by calendar period, reporting similar CD19^+^ counts, disease activity, EDSS, and adverse events over one year. Our within-patient switching design instead evaluates the same patients before and after transitioning from established originator treatment, in a younger, less disabled, mostly treatment-naive cohort (vs. 40.0% prior fingolimod, 27.5% natalizumab in Perez et al.), with longer follow-up (median 31.3 months vs. 1 year, including 18.9 months on biosimilar); no safety signal was identified on retrospective review despite adverse events not being systematically collected, and all patients persisted on therapy.

Rituximab remains the most cost-effective high-efficacy option by a wide margin at the drug-acquisition level, and the case for its broader adoption — including via biosimilar substitution — rests as much on this baseline affordability as on the additional savings biosimilar switching provides (Supplementary Table S5). This study has several design-related limitations: a single-center cohort, with some sub-analyses limited to small numbers of eligible patients (reported transparently with full denominators in the supplementary material); a retrospective, observational design with an unplanned “big-bang” switch that precludes a randomized comparison, though this reflects how nonmedical switches occur in real-world practice; and a switch implemented at the pharmacy level, with the treating physician becoming aware of it only retrospectively.

Adverse events were not systematically collected; complete persistence and the absence of a safety signal on retrospective review are reassuring but indirect tolerability indicators. Pre-rituximab radiological activity was defined by CELs alone — more lenient than the originator/biosimilar criterion, necessary given most patients lacked a comparable prior scan — likely underestimating true pre-rituximab activity; that the originator period still failed to separate from this conservative baseline reinforces our interpretation that depletion requires sustained exposure for full radiological effect.

These limitations are offset by methodological strengths: a within-patient design offering a more direct test of switching than the between-group comparison described above, with substantially longer follow-up, and an analysis combining paired and unpaired, patient- and scan-level approaches with appropriate correction for multiple comparisons. We believe these findings are worth reporting as, to our knowledge, the first description of clinical and radiological outcomes following a nonmedical switch from originator to biosimilar rituximab in pwMS.

## Conclusion

Nonmedical switching from originator to biosimilar rituximab was associated with sustained relapse suppression and progressive reduction in radiological activity, with comparable outcomes between products over a median follow-up of approximately two years. Complete treatment persistence further supports the tolerability of this switch in clinical practice. These findings support the use of rituximab in people with MS irrespective of which product is available. More broadly, real-world data should inform extrapolation decisions in similarly complex scenarios, improving adoption of rituximab as a cost-effective, high-efficacy MS therapy.

## Declarations

### Ethics Statement

This study was approved by the King Saud Medical City Institutional Review Board (approval number: H1RI-04-Apr23-04). Patient informed consent was not required, as data were collected exclusively from routine clinical records with no patient contact initiated for research purposes.

### Data Availability Statement

The dateset supporting the conclusions of this article is available from the corresponding author upon reasonable request and subject to institutional data governance approval.

### Author Contributions (CRediT Taxonomy)

Ahmed H. Althobaiti: Conceptualization, Data curation, Formal analysis, Inves-tigation, Methodology, Project administration, Supervision, Writing – original draft, Writing – review & editing Afnan A. Alnughaimish: Investigation, Writing – review & editing Saif S. Alqahtani: Data curation, resources, investigation, Data curation, Investigation, Writing – review & editing Fahad Aldosarri: Data curation, Investigation, Writing – review & editing

## Acknowledgements

The authors would like to thank the staff of King Saud Medical City (KSMC), Riyadh, Saudi Arabia, for their support in facilitating access to the medical records used in this study, and the staff of the Riyadh Regional Laboratory, Department of Immunology and Flow Cytometry, for their contribution to the laboratory analyses supporting this work. Artificial intelligence (Claude, Anthropic) was used solely for language editing and rephrasing during manuscript preparation. The authors are responsible for all scientific content, data, analyses, interpretations, and conclusions in this manuscript, and reviewed and approved the final version.

---

**Supplementary Figure 1.**
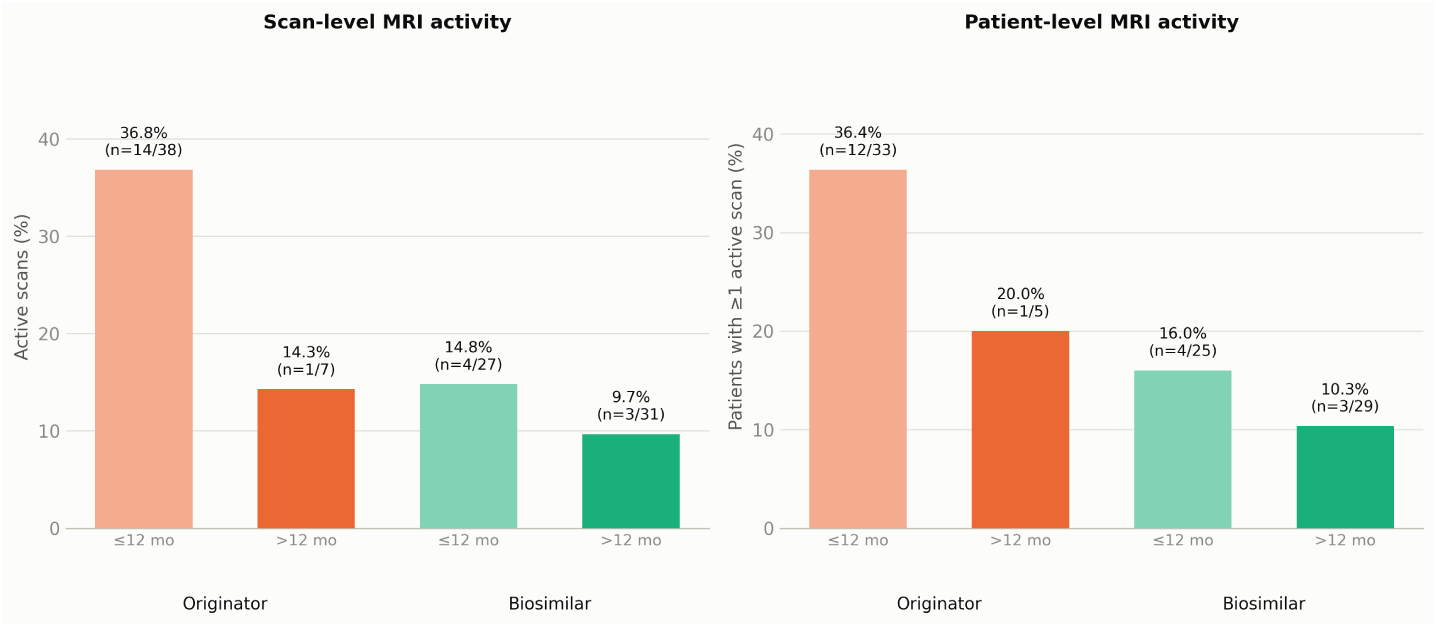
MRI activity by product and time since initiation.

**Supplementary Figure 2.**
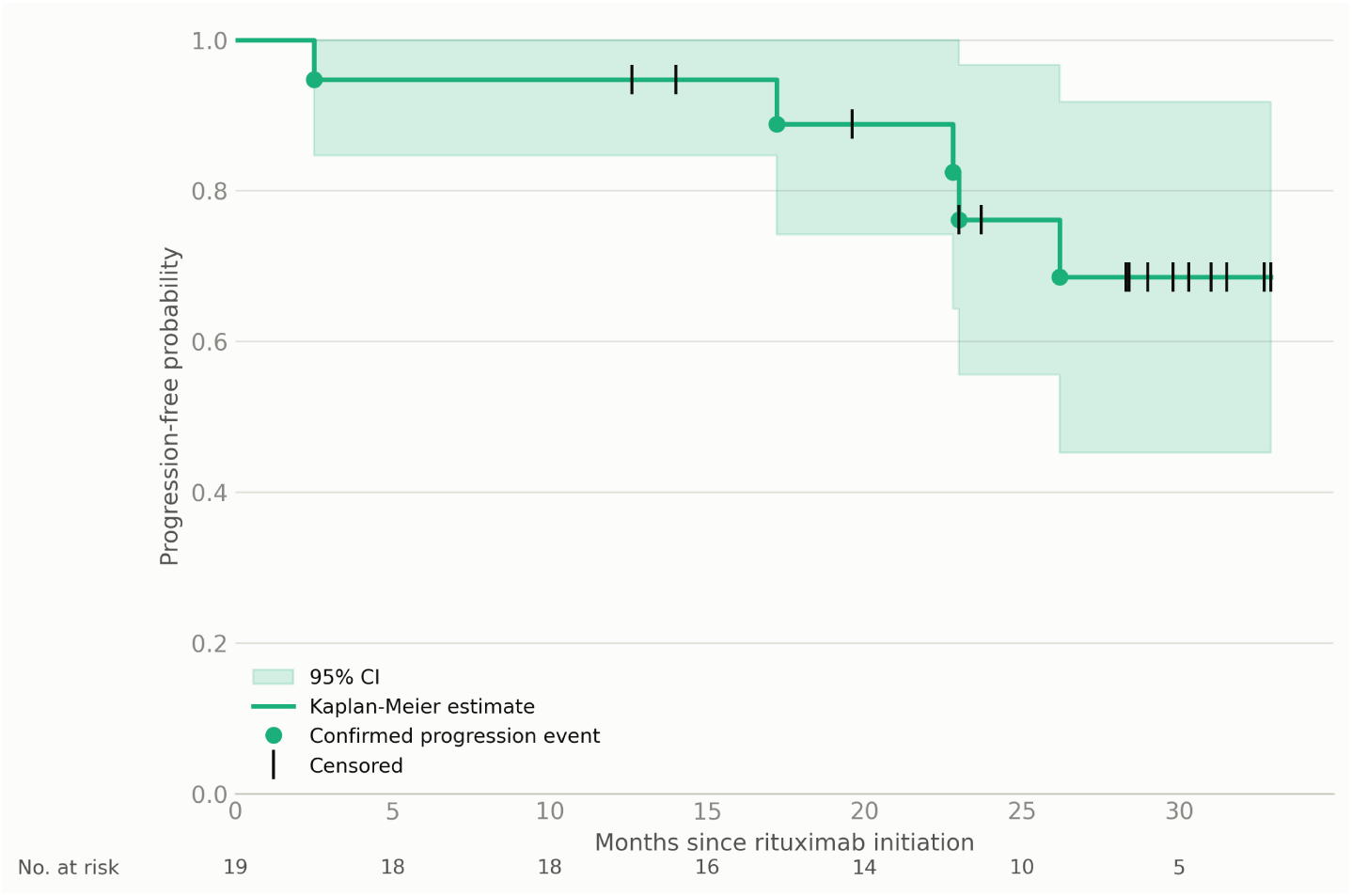
EDSS trajectories — confirmed progressors (n=5).

**Supplementary Figure 3.**
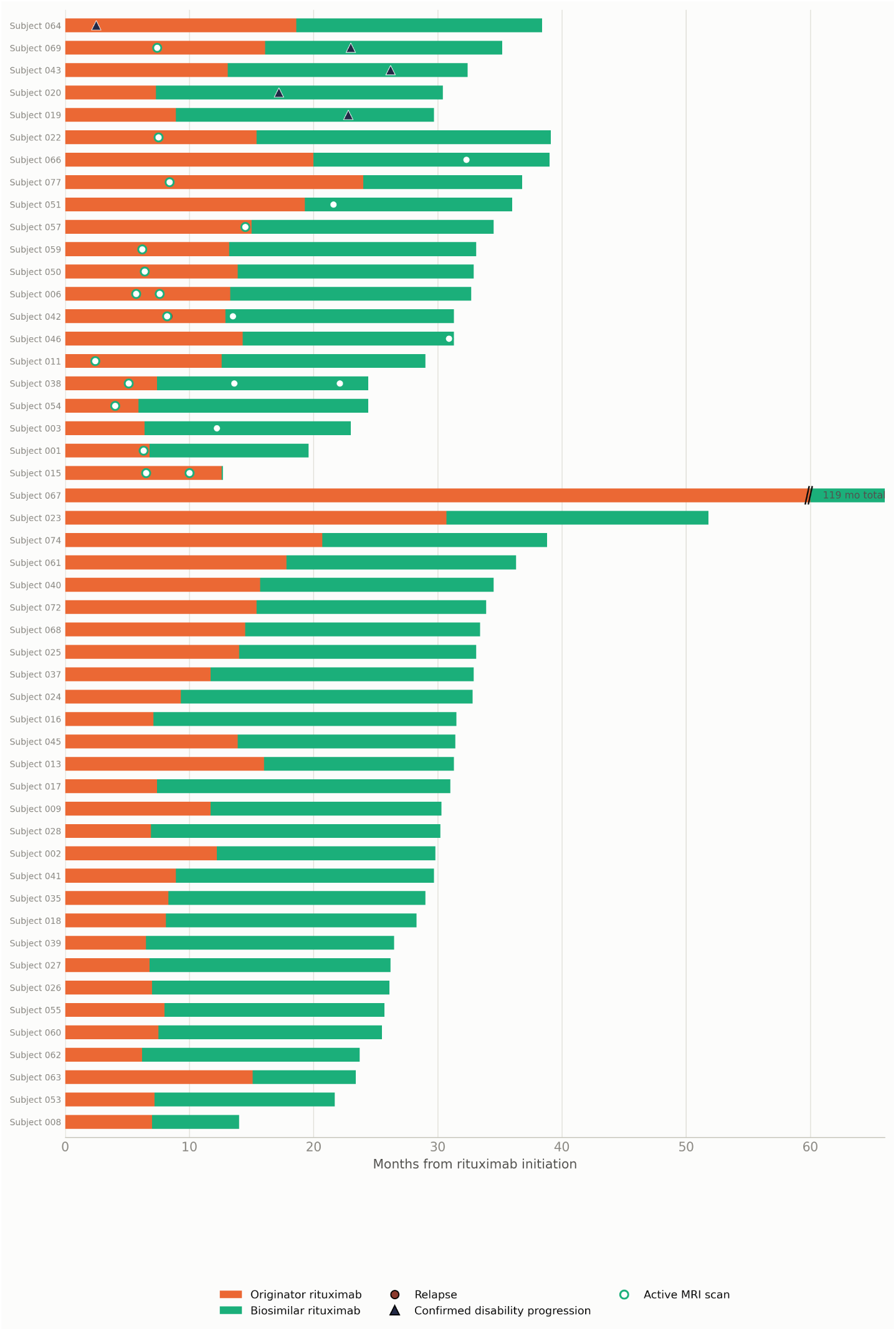
Patient-level treatment timeline: originator-to-biosimilar rituximab switch. Axis capped at 60 months; bars extending beyond are marked with a break and labeled with total duration. Sorted: confirmed progressors, then patients with a relapse or active scan, then remaining patients by descending treatment duration.

**Supplementary Figure 4.**
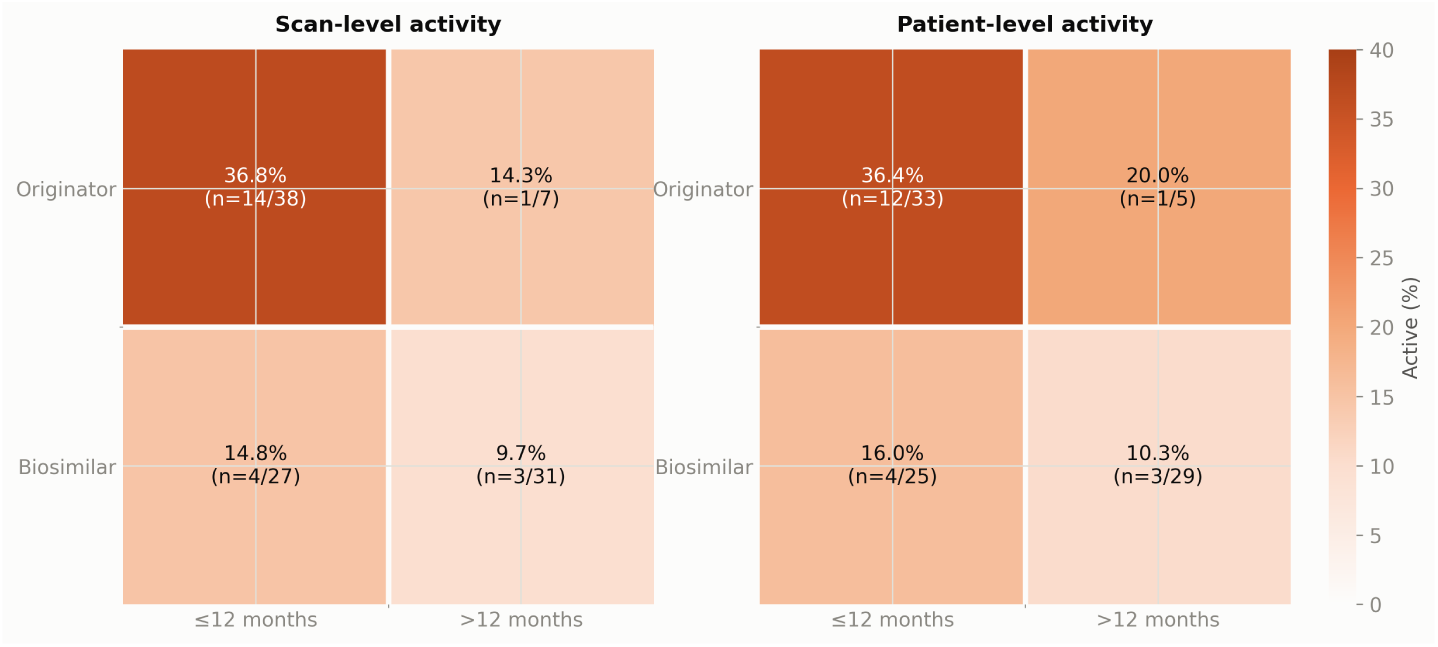
MRI activity by product and time since initiation.

**Supplementary Figure 5.**
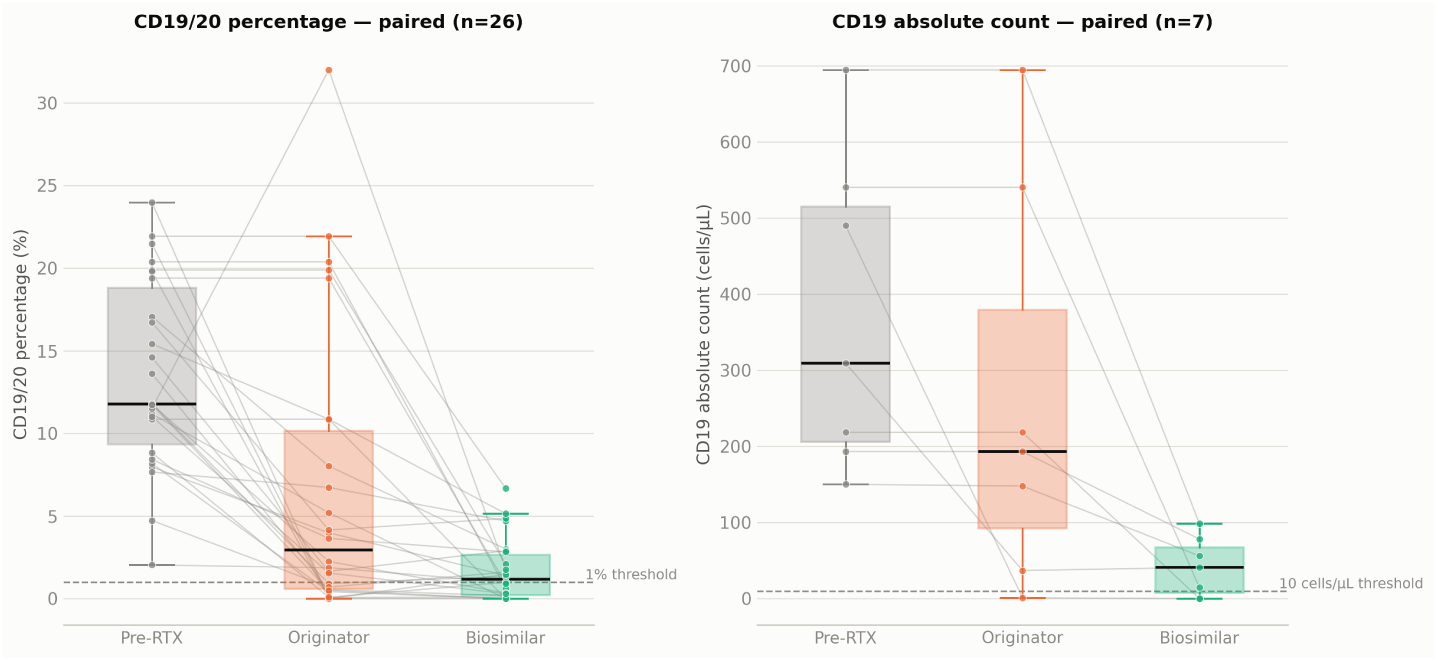
CD19/20 B-cell measures across treatment eras — paired patients.

**Supplementary Figure 6.**
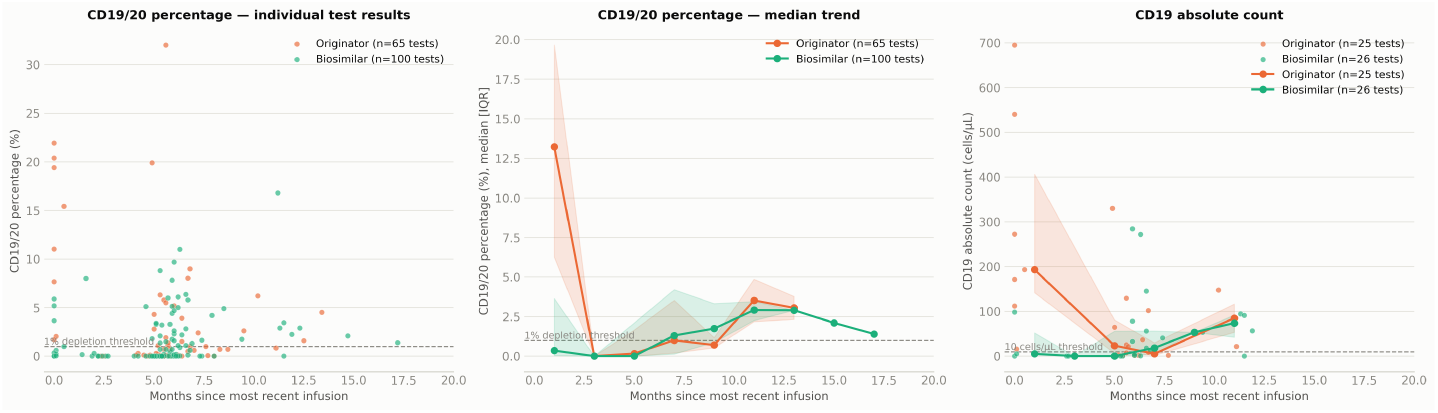
CD19/20 B-cell measures across treatment eras — paired patients. Each line connects one patient’s pre-RTX value, median-on-originator value, and median-on-biosimilar value. The absolute-count panel reflects a small paired sample and should be interpreted with caution.

**Supplementary Figure 7.**
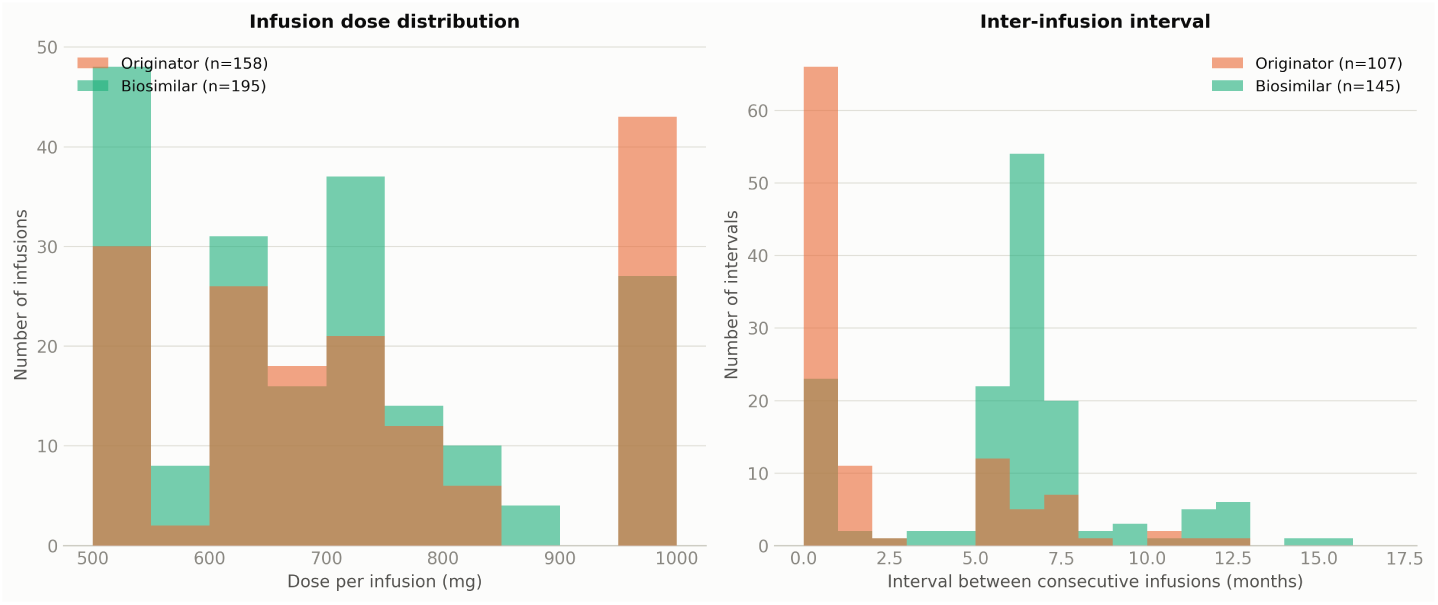
Rituximab dosing and infusion timing: originator vs biosimilar. 1 originator-period interval(s) beyond 16 months (treatment gap, single patient) excluded from the interval panel for readability.

**Supplementary Figure 8.**
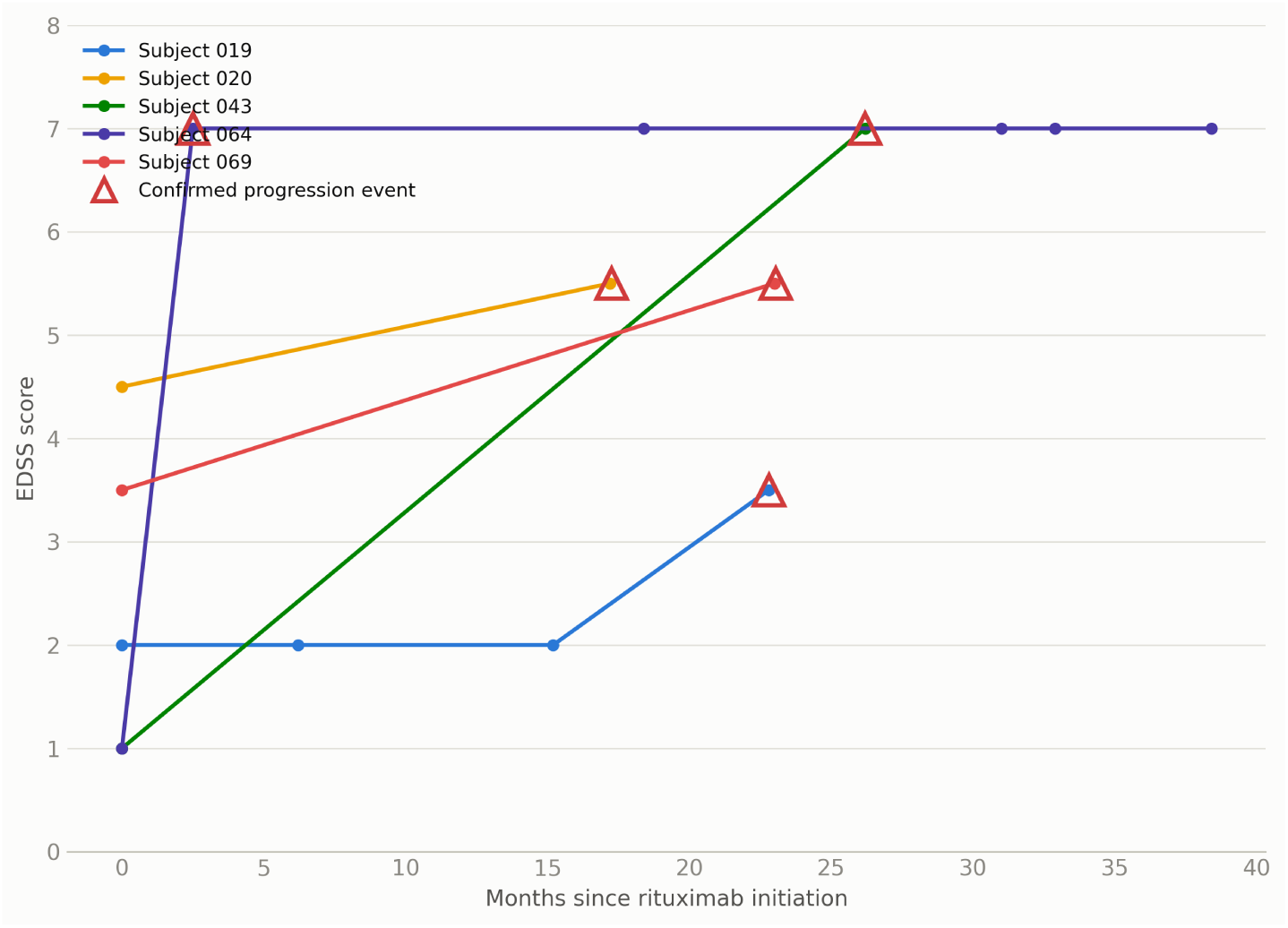
EDSS trajectories — confirmed progressors (n=5).

**Supplementary Figure 9.**
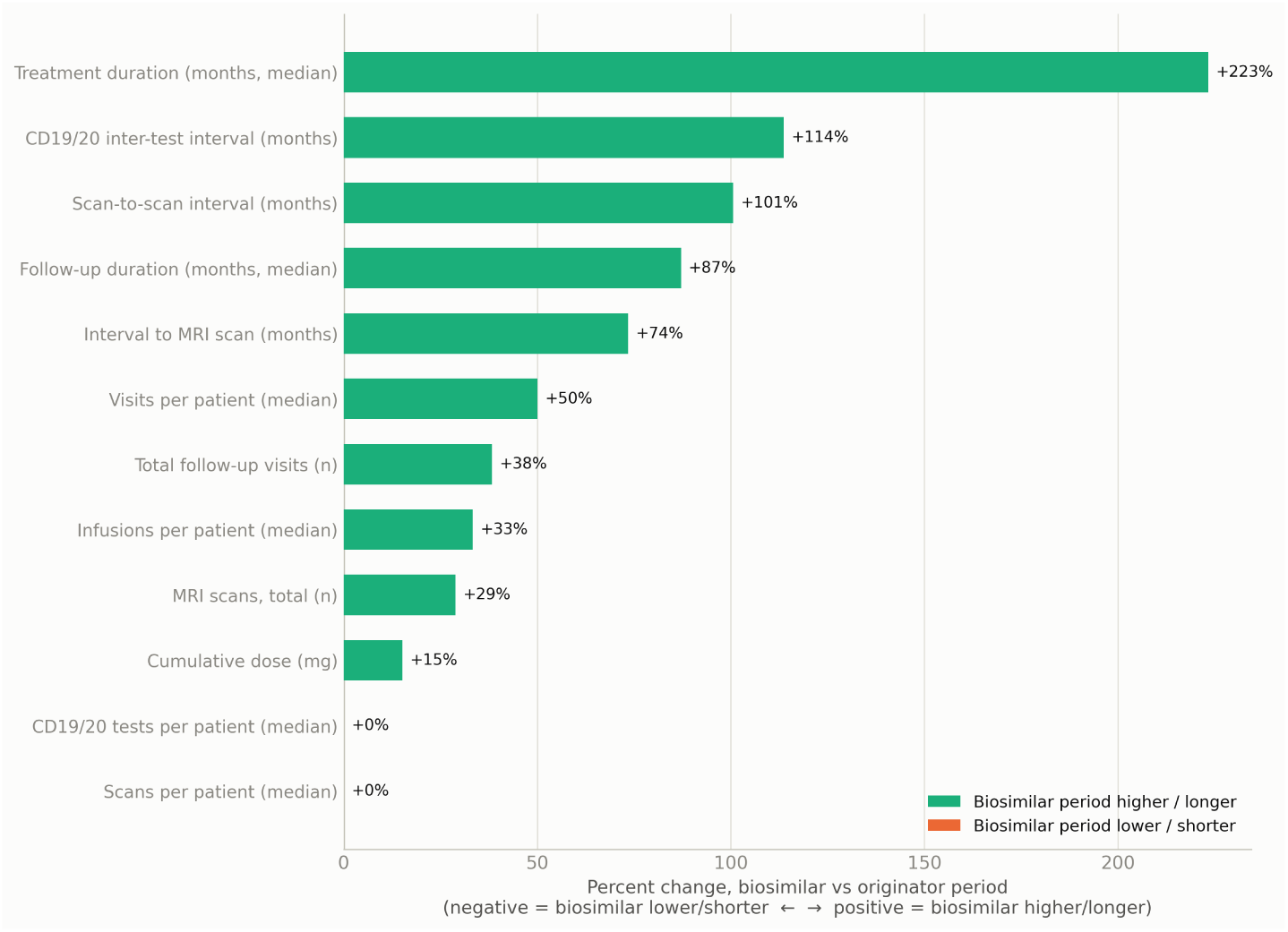
Data collection asymmetry: originator vs biosimilar treatment period. Reflects unequal observation windows, not necessarily true clinical differences: the biosimilar period generally had longer follow-up and more monitoring per patient.

**Supplementary Figure 10.**
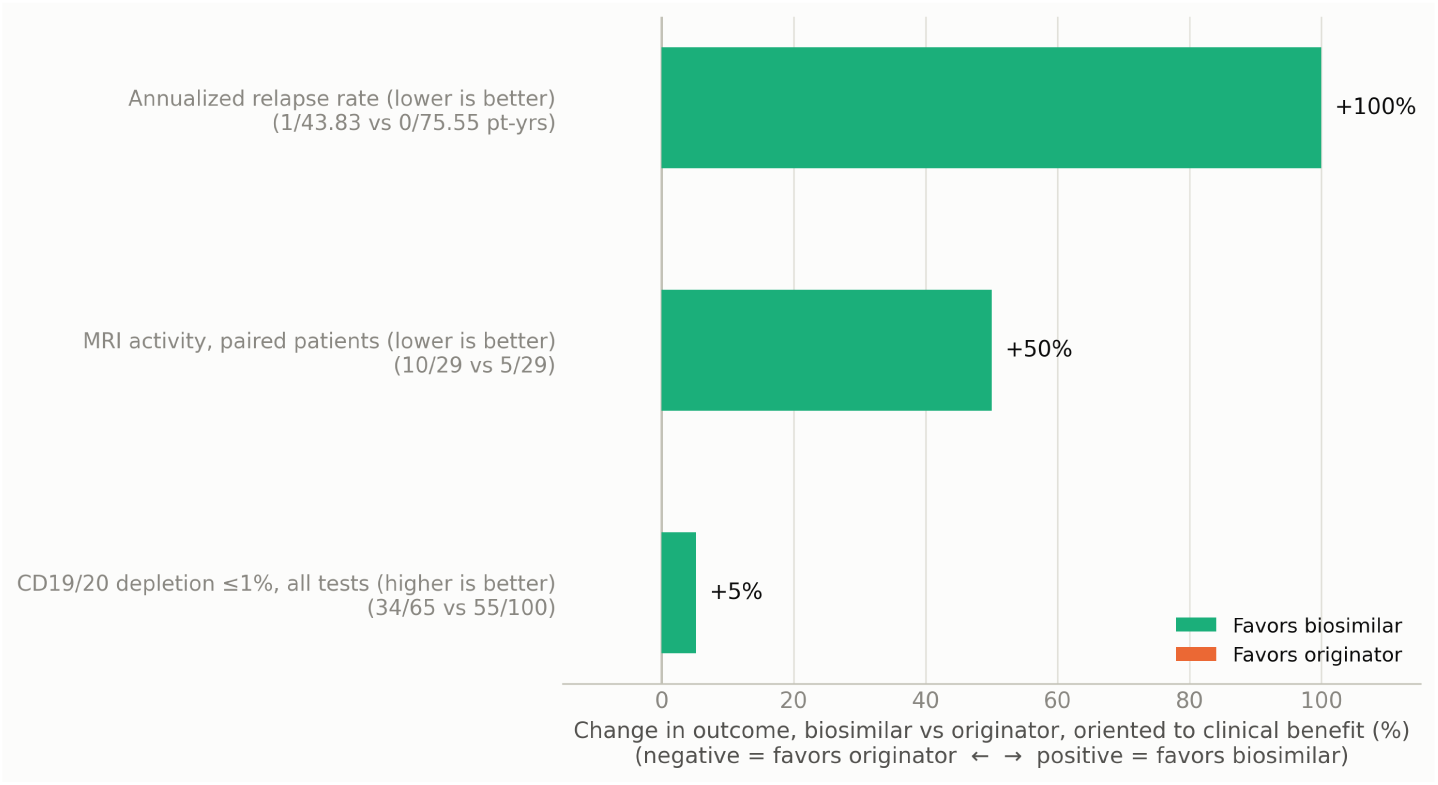
Clinical outcome differences: originator vs biosimilar rituximab. ARR comparison reflects 0 relapses observed on biosimilar (descriptive only — see Figure 2 for full CIs). None of these differences reached statistical significance (see Figures 1 and 2 for confidence intervals).

**Supplementary Figure 11.**
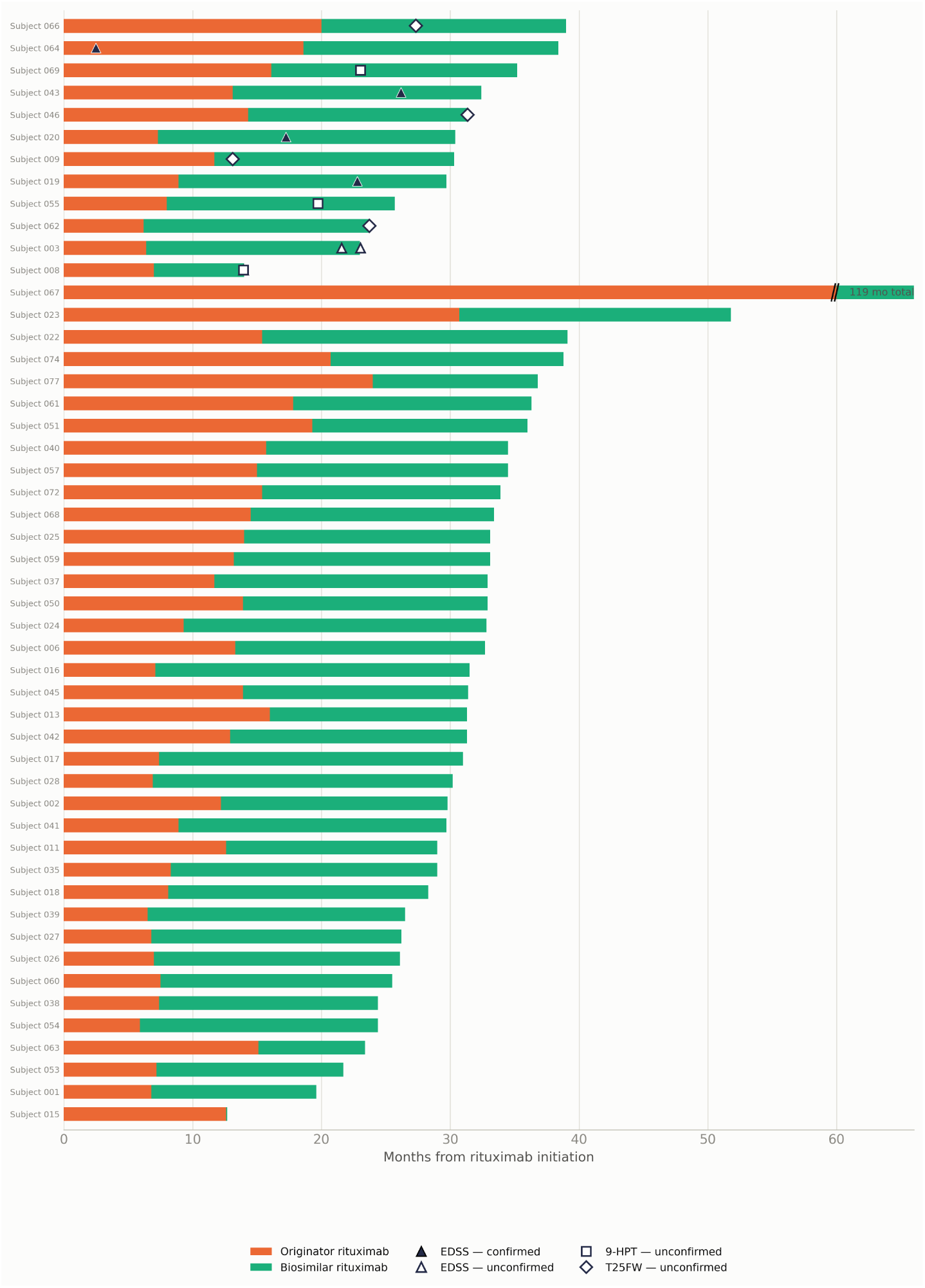
Functional progression events across all domains (n=50). Solid markers: confirmed progression (180-day confirmation). Hollow markers: unconfirmed events (no qualifying confirmation visit available). 9-HPT combines right- and left-hand tests.

**Supplementary Table S1.** Eligibility for Disability Progression Analysis.

|  | EDSS | NHPT | T25FW |
| --- | --- | --- | --- |
| Data available at baseline (pre-rituximab) | 43 | 7 | 11 |
| Patients with 1 follow-up score | 20 | 46 | 41 |
| Eligible (2 scores 6 months apart) | 19 | 17 | 21 |
| Not eligible (insufficient follow-up) | 24 | 33 | 29 |

EDSS = Expanded Disability Status Scale; NHPT = Nine-Hole Peg Test; T25FW = Timed 25-Foot Walk.
Eligibility requires: (1) a baseline score (from the baseline visit or first available follow-up visit); and (2) at least two additional follow-up assessments separated by a minimum of 6 months. NHPT and T25FW did not meet the 60% data-availability threshold for primary reporting and are presented in supplementary material only.

**Supplementary Table S2.** Data Collection Asymmetry Between Origi-nator and Biosimilar Periods.

| Variable | Originator period | Biosimilar period |
| --- | --- | --- |
| **Clinical follow-up** |  |  |
| Total follow-up visits | 101 | 154 |
| Patients with no follow-up visit | 11 (22.0%) | 1 (2.0%) |
| Visits per patient | 2 (IQR 2–4, range 1–6) | 3 (IQR 2–4, range 1–9) |
| Median follow-up duration, months | 9.0 (IQR 6.9–15.0, range 2–102.8) | 18.9 (IQR 17.5–20.0, range 7–24.4) |
| **MRI** |  |  |
| Total scans | 45 | 58 |
| Patients with no MRI | 14 (28.0%) | 10 (20.0%) |
| Scans per patient | 1 (IQR 1–1, range 1–3) | 1 (IQR 1–2, range 1–3) |
| Median interval from treatment initiation to scan, months | 7.1 (IQR 6.3–10.0, range 2.4–25.3) | 12.0 ± 6.8 (mean ± SD, range 0.6–25.0) |
| Mean scan-to-scan interval, months | 5.9 ± 3.7 | 10.9 ± 4.1 |
| Scans acquired within 12 months of initiation | 38/45 (84.4%) | 27/58 (46.6%) |
| **Rituximab infusions** |  |  |
| Infusions per patient | 3 (IQR 3–4, range 2–5) | 4 (IQR 4–4, range 1–6) |
| Total cumulative dose, mg | 2,050 (IQR 1,825–2,775, range 1,200–4,000) | 2,648.1 ± 795.2 (mean ± SD, range 600–4,300) |
| Treatment duration, months | 6.0 (IQR 0.9–8.0, range 0.5–96.3) | 19.4 (IQR 16.1–20.1, range 0.5–24.4) |
| **CD19/CD20 laboratory tests** |  |  |
| Patients tested | 42 (84.0%) | 44 (88.0%) |
| Tests per patient | 2 (IQR 1–2, range 1–6) | 2 (IQR 2–3, range 1–5) |
| Median inter-test interval, months | 2.9 (IQR 2–7, range 0–14.6) | 2.0 (IQR 2–3, range 1–5) |

Data are median (IQR) or mean ± SD unless otherwise stated.
Treatment duration excludes 2 patients with a single biosimilar infusion.

Supplementary Tables S3-S4. MRI activity stratified by time (12 months vs >12 months)

**Table S3.** MRI activity — patient level, unpaired (full cohort)

| Sub-period | Patients | Active patients | Active (%) |
| --- | --- | --- | --- |
| Originator 12m | 33 | 12 | 36.4% |
| Originator >12m | 5 | 1 | 20.0% |
| Biosimilar 12m | 25 | 4 | 16.0% |
| Biosimilar >12m | 29 | 3 | 10.3% |

**Table S4.**
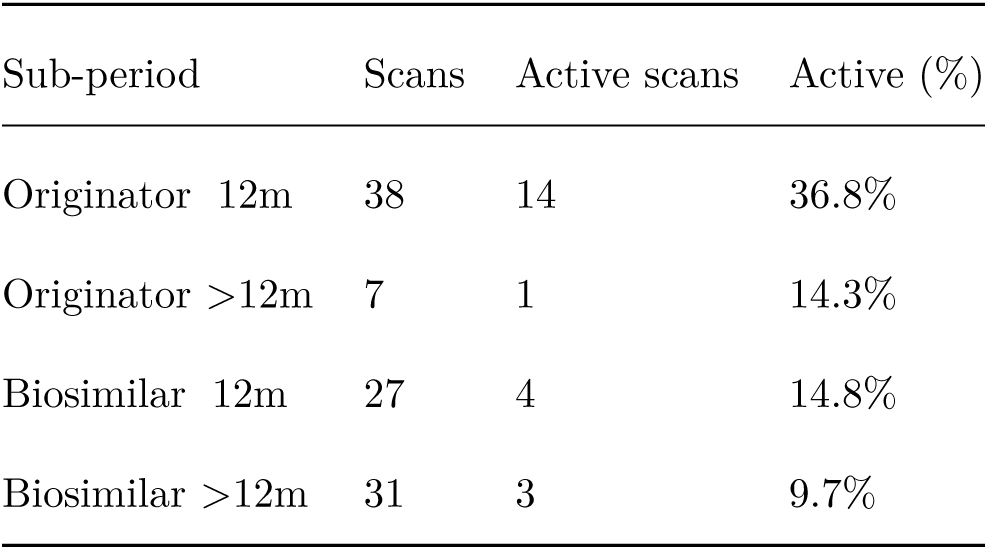
MRI activity — scan level, unpaired (full cohort)

| Sub-period |  | Scans | Active scans | Active (%) |
| --- | --- | --- | --- | --- |
| Originator | 12m | 38 | 14 | 36.8% |
| Originator | >12m | 7 | 1 | 14.3% |
| Biosimilar | 12m | 27 | 4 | 14.8% |
| Biosimilar | >12m | 31 | 3 | 9.7% |

**Supplementary Table S5.**
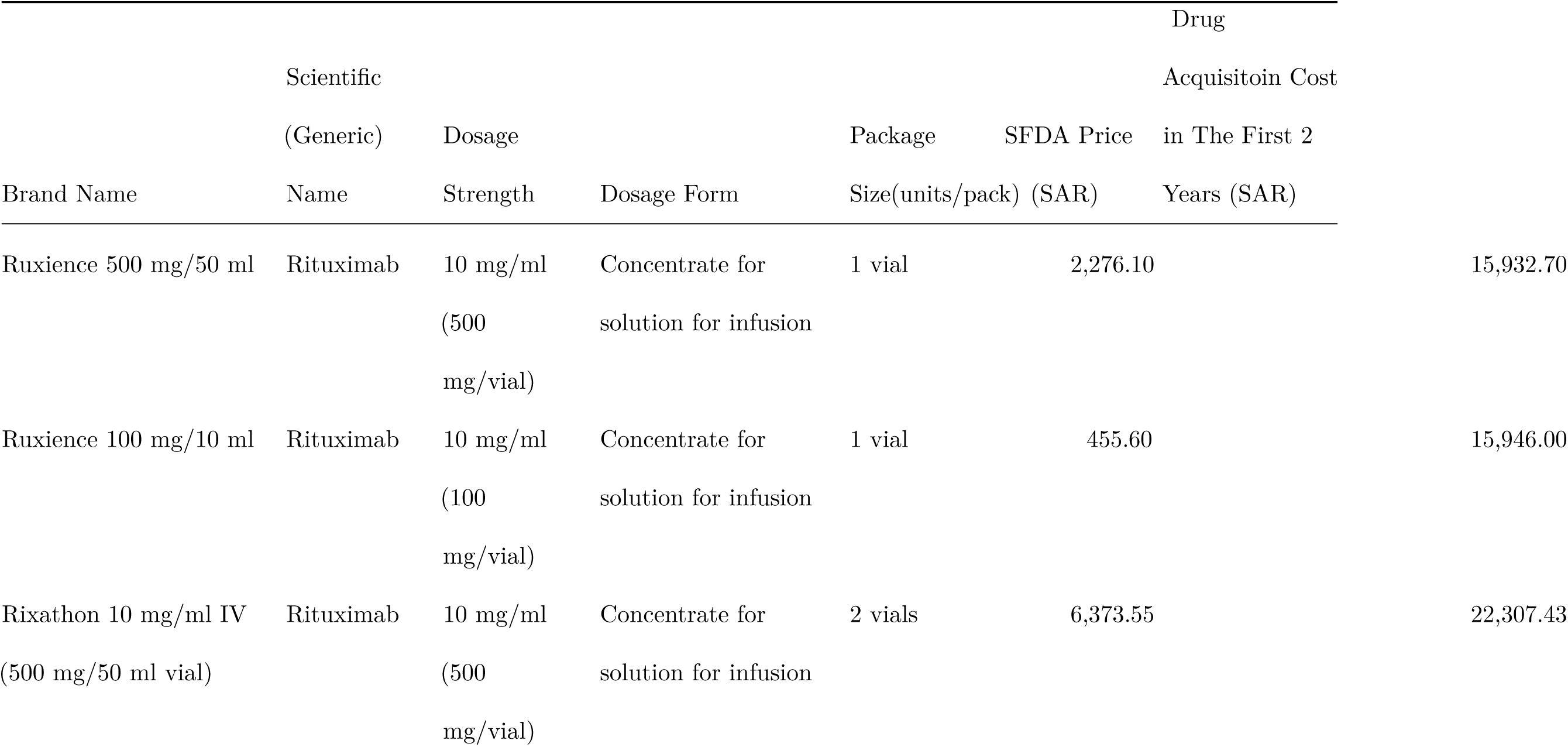
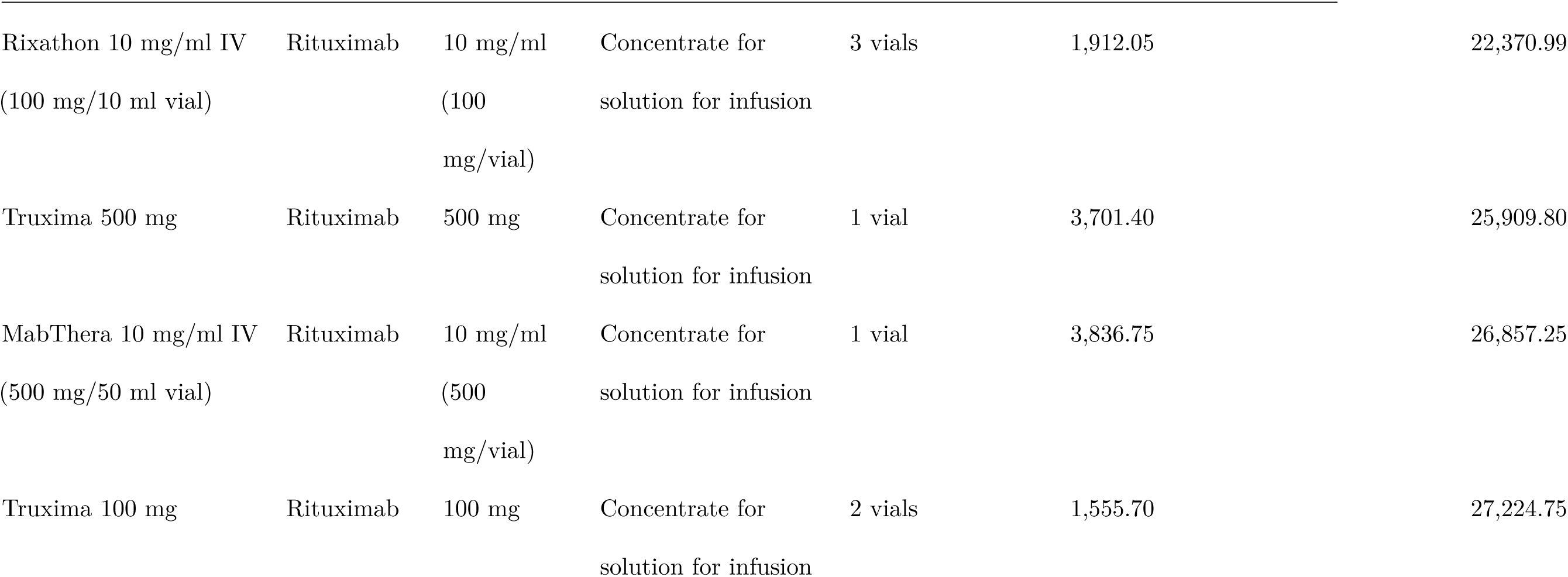
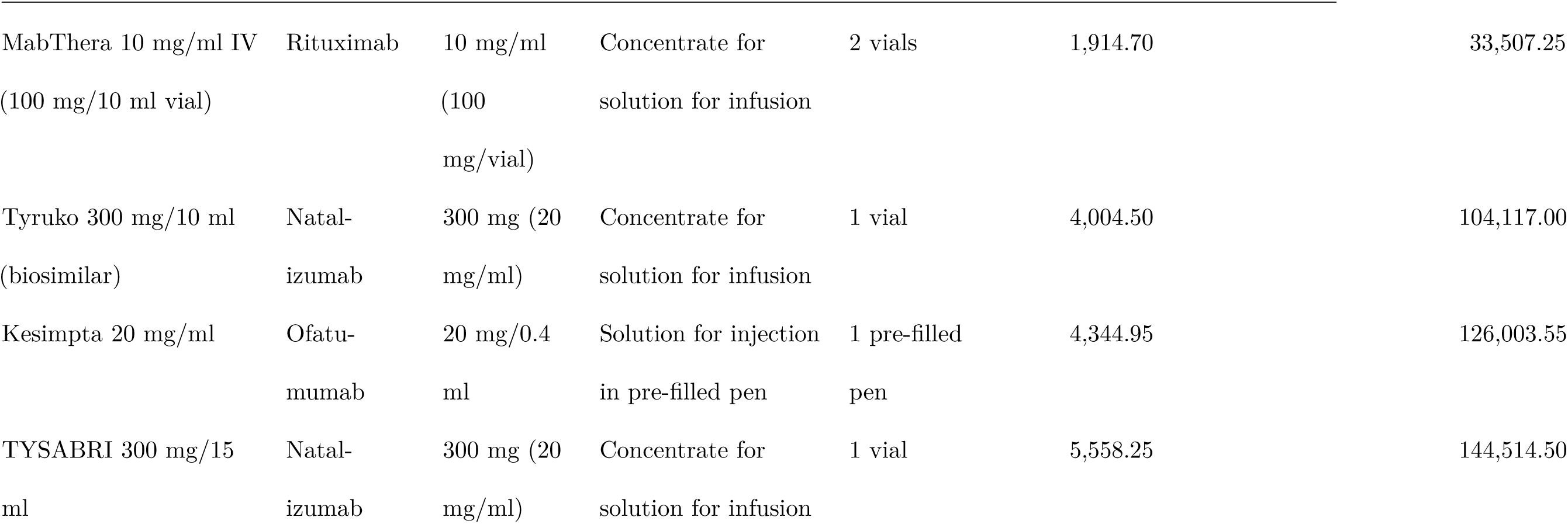
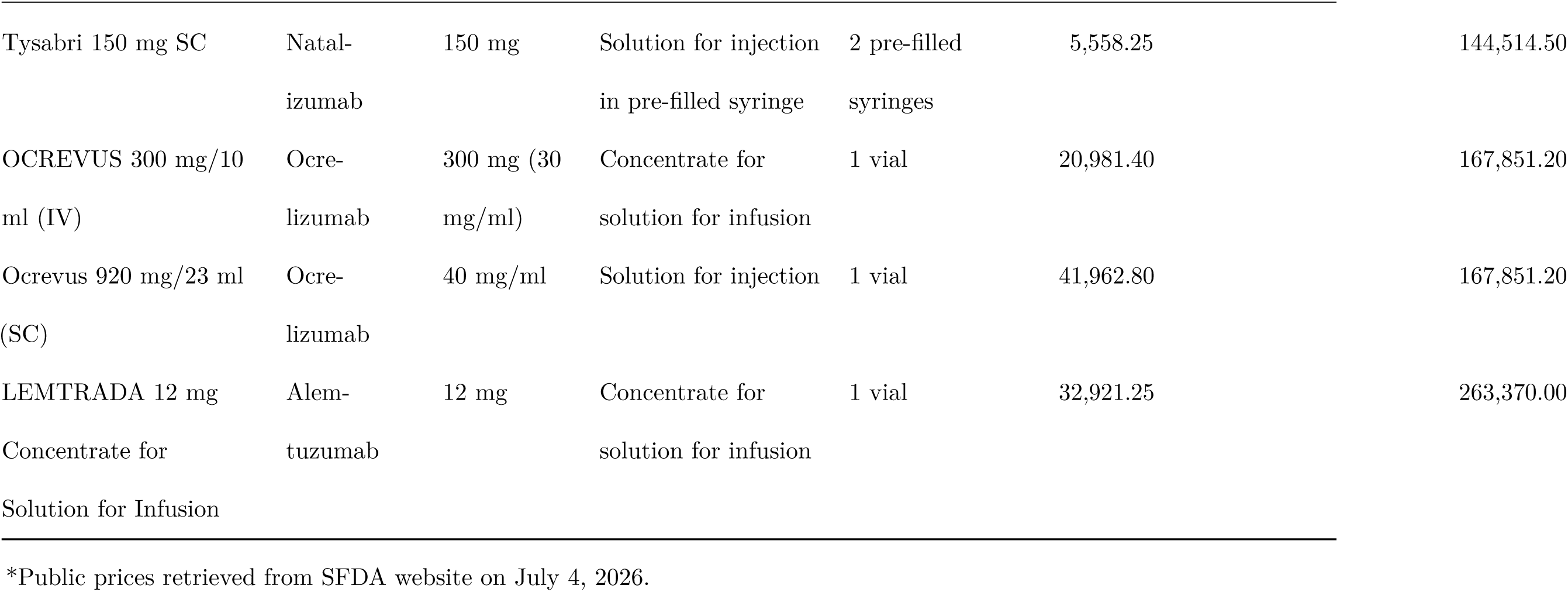
Highly Effective Therapy Drug Acquisition Cost Over First 2 Years of Treatment.

