## supplementary material for "Effectiveness and Tolerability of Nonmedical Switching from Originator (MabThera®) to Biosimilar (Truxima®) Rituximab in People with Multiple Sclerosis: A Tertiary Single-Center Observational Study"

### 1 Supplementary Material

#### 1.1 Supplementary Table S1. Eligibility for Disability Progression Analysis

`Supplementary Table S1 here.`

---

#### 1.2 Supplementary Table S2. Data Collection Asymmetry Between Originator and Biosimilar Periods

Substantial differences in data collection were observed between the originator and biosimilar treatment periods (Supplementary Table S2).

`Supplementary Table S2 here.`

Patients had fewer total follow-up visits during the originator period (101 visits among 39 patients, with 11 patients having no follow-up visit) compared with the biosimilar period (154 visits among 49 patients, with 1 patient having no follow-up visit). The median number of visits per patient was 2 (IQR 2–4, range 1–6) during the originator period and 3 (IQR 2–4, range 1–9) during the biosimilar period. Median clinical follow-up duration was shorter during the originator period (9 months, IQR 6.9–15.0, range 2–102.8) than during the biosimilar period (18.9 months, IQR 17.5–20.0, range 7–24.4).

Patients underwent fewer MRI scans during the originator period (45 scans among 36 patients, with 14 patients having no MRI) than during the biosimilar period (58 scans among 40 patients, with 10 patients having no MRI). The median number of scans per patient was 1 (IQR 1–1, range 1–3) during the originator period and 1 (IQR 1–2, range 1–3) during the biosimilar period. The median interval from treatment initiation to scan was shorter during the originator period (7.1 months, IQR 6.3–10.0, range 2.4–25.3) than during the biosimilar period (mean 12.0 ± 6.8 months, range 0.6–25.0). The mean scan-to-scan interval was 5.9 ± 3.7 months during the originator period and 10.9 ± 4.1 months during the biosimilar period. Thirty-eight of 45 originator-period scans were obtained within 12 months of treatment initiation, compared with 27 of 58 biosimilar-period scans; conversely, 7 of 45 originator-period scans and 31 of 58 biosimilar-period scans were obtained 12 months or more after treatment initiation.

Patients received fewer rituximab infusions during the originator period (median 3, IQR 3–4, range 2–5) than during the biosimilar period (median 4, IQR 4–4, range 1–6), and a lower total cumulative dose (median 2,050 mg, IQR 1,825–2,775, range 1,200–4,000) than during the biosimilar period (mean 2,648.1 ± 795.2 mg, range 600–4,300). Median treatment duration was shorter during the originator period (6 months, IQR 0.9–8.0, range 0.5–96.3) than during the biosimilar period (19.4 months, IQR 16.1–20.1, range 0.5–24.4; two patients with a single infusion were excluded from this analysis).

The median number of tests per patient was 2 (IQR 1–2, range 1–6) during the originator period and 2 (IQR 2–3, range 1–5) during the biosimilar period. The median inter-test interval was 2.9 months (IQR 1.5–7.2) during the originator period and 6.2 months during the biosimilar period (mean 5.9 ± 2.5 months); this difference did not reach statistical significance (Mann–Whitney U = 793.0, p = 0.086).

---

#### 1.3 Supplementary Results: Scan Timing and Interval Data

Detailed scan timing data for the Radiological outcomes section.

Among all rituximab-era scans, the median number of scans per patient was 2 (IQR 2–3, range 1–5). During the originator period, the median interval from treatment initiation to scan was 7.1 months (IQR 6.3–10.0, range 2.4–25.3), with a median of 1 scan per patient (IQR 1–1). During the biosimilar period, the mean interval from treatment initiation to scan was 12.0 ± 6.8 months (range 0.6–25.0), with a median of 1 scan per patient (IQR 1–2). The majority of originator-period scans (38 of 45, 84.4%) were acquired within the first 12 months of treatment, compared with fewer than half of biosimilar-period scans (27 of 58, 46.6%).

---

#### 1.4 Supplementary Results: Scan Timing Comparability and Within-Biosimilar-Period Analysis

Scan timing was comparable between the beyond-12-month originator and biosimilar subgroups, with mean intervals from treatment initiation of 15.2 ± 2.6 months for the originator period and 17.6 ± 3.0 months for the biosimilar period (independent t-test, $p = 0.098$). Within the biosimilar period, activity did not differ significantly between scans obtained within 12 months and those obtained beyond 12 months, either by patient-level binary classification (4 of 25 vs. 3 of 29; Fisher exact, $p = 0.692$) or by active-scan proportion (Wilcoxon, $p = 0.180$).

---

#### 1.5 Supplementary Results: Disability Progression

##### 1.5.1 EDSS

Among the 19 patients eligible for EDSS progression analysis, 5 (26.3%) had a confirmed disability progression event: 4 during biosimilar treatment and 1 during originator treatment. Three additional EDSS worsening events occurred during biosimilar treatment but were not confirmed — one had insufficient follow-up (<180 days) to assess confirmation, and two had no subsequent visit.

`Supplmentary Figure 2: EDSS trajectories - confirmed progressors (n=5).`

##### 1.5.2 9-Hole Peg Test (NHPT)

Among the 17 patients eligible for 9-HPT progression analysis, no confirmed progression events occurred in either hand. Three unconfirmed worsening events were observed in the right hand, all during biosimilar treatment: two had no subsequent visit, and one had recovered by the confirmation visit. No worsening events, confirmed or unconfirmed, were observed in the left hand.

##### 1.5.3 Timed 25-Foot Walk (T25FW)

Among the 21 patients eligible for T25FW progression analysis, no confirmed progression events occurred. Four unconfirmed worsening events were observed, all during biosimilar treatment and all lacking a subsequent visit for confirmation.

`Supplementary Figure 3: Patient-level treatment timeline: originator-to-biosimilar rituximab switch.`

---

#### 1.6 Supplementary Results: Stratification of radiological activity by time ( 12 months or less vs more than 12 months)

Most originator-period radiological activity occurred within the first 12 months of treatment (12 active patients within 12 months versus 1 beyond 12 months). Among scans showing activity during the originator period within the first 12 months, the mean interval from treatment initiation to the active scan was 6.6

± 1.8 months. Activity during the biosimilar period continued to decline both within and beyond 12 months (4 of 25 patients, 16.0%, versus 3 of 29 patients, 10.3%).

The proportion of active patients did not differ significantly between scans obtained 12 months or more after originator initiation and those obtained 12 months or more after biosimilar initiation (20.0% vs. 10.3%, p = 0.488).

`Supplementary Figure 4. MRI activity by product and time since initiation.`

Scan timing was comparable between the beyond-12-month originator and biosimilar subgroups, and activity within the biosimilar period did not differ significantly by scan timing; full results are reported in the supplementary material. (Supplementary Table S2)

#### 1.7 Supplementary Results: Paired Radiological Analysis Beyond 12 Months

Paired analysis restricted to patients with both originator-period and biosimilar-period scans obtained more than 12 months after treatment initiation was limited to 5 patients. Given this sample size, results are reported descriptively only and should be interpreted with extreme caution: of these 5 patients, 1 of 5 (20.0%) showed activity on the originator-period beyond-12-month scan and 0 of 5 (0.0%) showed activity on the biosimilar-period beyond-12-month scan.

#### 1.8 Supplementary Methods: Laboratory Assay Protocols

##### 1.8.1 1. Lymphocyte subset immunophenotyping by BD Multitest 6-Color TBNK with BD Trucount tubes

Peripheral blood lymphocyte subsets were analyzed by flow cytometry using the BD Multitest 6-Color TBNK reagent with BD Trucount tubes (BD Biosciences, San Jose, CA, USA) on a BD FACSCanto II flow cytometer. The assay was used to determine the percentages and absolute counts of mature lymphocyte subsets, including CD3+ T lymphocytes, CD19+ B lymphocytes, CD3-CD16+/CD56+ natural killer (NK) cells, CD3+CD4+ helper T cells, and CD3+CD8+ cytotoxic T cells.

**Specimen type and sample handling**

Fresh peripheral whole blood collected by venipuncture into EDTA anticoagulant tubes was used for analysis. Samples were maintained at room temperature (20–25°C) and stained within 24 hours of collection. Stained preparations were acquired within 6 hours after staining.

**Staining procedure**

For absolute counting, staining was performed directly in BD Trucount tubes according to the manufacturer's instructions. Briefly, 20 µL of BD Multitest 6-Color TBNK reagent was added to the tube, followed by 50 µL of well-mixed anticoagulated whole blood. The blood specimen was added carefully using a reverse pipetting technique to improve volume accuracy and avoid loss of material along the tube wall. Tubes were gently vortexed and incubated for 15–30 minutes at room temperature in the dark. After incubation, 450 µL of 1× BD FACS Lysing Solution was added, the tubes were mixed gently, and incubated again for 15–30 minutes in the dark at room temperature. There was no wash step. Samples were vortexed gently immediately before acquisition.

**Instrument setup and compensation**

Acquisition was performed on a BD FACSCanto II flow cytometer using the recommended clinical setup for this platform. The instrument underwent daily quality control before patient testing. Detector settings and instrument sensitivity were verified using the appropriate BD setup beads for the FACSCanto II platform. Fluorescence compensation was established using the BD FACS 7-Color Setup Beads and instrument-associated clinical software, in accordance with the manufacturer's recommendations. The acquisition threshold was adjusted to minimize debris while maintaining inclusion of the lymphocyte population and Trucount bead events.

**Quality control**

Daily internal quality control was performed before analysis of patient samples using two levels of controls: BD Multi-Check Control and BD Multi-Check CD4 Low Control. Controls were stained and processed in the same manner as patient samples and acquired on the same instrument. Control results were accepted only when values fell within the manufacturer-assigned ranges, and the CD45 versus side scatter (SSC) distribution showed appropriate separation of lymphocytes, monocytes, and granulocytes.

##### Data acquisition and analysis

Data were acquired and analyzed using cytometer-specific BD clinical analysis software. Lymphocytes were identified primarily by CD45-bright/low SSC characteristics. The reagent system uses fluorescence-assisted gating to improve resolution of the lymphocyte population and reduce contamination by debris, unlysed erythrocytes, or non-lymphoid events. Trucount bead events were identified separately, and absolute cell counts (cells/µL) were calculated using the single-platform bead-based method by comparing cellular events with bead events. The following subsets were reported as both percentage of lymphocytes and absolute count: CD3+, CD19+, CD3-CD16+/CD56+, CD3+CD4+, and CD3+CD8+ cells. All dot plots were visually reviewed to confirm correct gating.

3. Clinical and Laboratory Standards Institute. Enumeration of Immunologically Defined Cell Populations by Flow Cytometry; Approved Guideline. CLSI document H42. Wayne, PA: CLSI.

---

###### 1.8.2 2. CD19/CD20 screening by flow cytometry

A semi-quantitative CD19/CD20 screening assay was performed on a BD FACSCanto II flow cytometer, and data were analyzed using FACSDiva software (BD Biosciences, San Jose, CA, USA). The assay utilized CD45 PerCP, CD20 FITC, and CD19 APC monoclonal antibodies (BD Biosciences, San Jose, CA, USA).

**Specimen type and sample handling**

The assay was performed on fresh peripheral blood collected into EDTA tubes. Samples were kept at room temperature and processed within 24 hours of collection.

**Staining and sample preparation**

Briefly, 100 µL of peripheral blood was added to a 12 × 75 mm tube containing 20 µL of CD45 PerCP, 20 µL of CD20 FITC, and 5 µL of CD19 APC. The tube was vortexed vigorously for 7–10 seconds and incubated for 20 minutes at room temperature in the dark. Subsequently, 2 mL of 1× BD FACS Lysing Solution was added, followed by incubation for 10–15 minutes at room temperature in the dark. The preparation was then centrifuged for 5 minutes at 300 rpm, and the supernatant was discarded. The cell pellet was washed with 2 mL of cell wash, centrifuged again for 5 minutes at 300 rpm, and the supernatant was discarded. Finally, the cells were resuspended in 0.2 mL of cell wash, vortexed briefly, and acquired immediately.

##### Instrument setup, compensation, and quality assurance

The BD FACSCanto II was subjected to routine daily instrument setup and performance verification before sample acquisition. Daily fluorescence compensation was established before testing. In addition, a fresh peripheral blood sample from a healthy donor was processed and acquired with each patient sample as an in-run biological reference to verify expected staining pattern and antigen distribution.

##### Data acquisition and analysis

Analysis was performed using a sequential gating strategy in FACSDiva software. Leukocytes were first identified, and lymphocytes were gated based on CD45 expression and side scatter characteristics. Within the lymphocyte gate, CD19- and CD20-positive B-cell populations were assessed, and results were expressed as the percentage of gated lymphocytes showing CD19 and/or CD20 expression, according to the laboratory's semi-quantitative reporting approach. Dot plots were visually inspected to confirm adequate separation of positive and negative populations and overall technical acceptability.

2. Clinical and Laboratory Standards Institute. Enumeration of Immunologi-

cally Defined Cell Populations by Flow Cytometry; Approved Guideline. CLSI document H42. Wayne, PA: CLSI.

3. Craig FE, Foon KA. Flow cytometric immunophenotyping for hematologic neoplasms. Blood. 2008;111(8):3941–3967.
